# Quantifying threat from COVID-19 infection hazard in Primary Schools in England

**DOI:** 10.1101/2020.08.07.20170035

**Authors:** W.P. Aspinall, R.S.J. Sparks, R.M. Cooke, J. Scarrow

## Abstract

We have constructed a COVID-19 infection hazard model for the return of pupils to the 16,769 state Primary Schools in England that takes into account uncertainties in model input parameters. The basic probabilistic model estimates likely number of primary schools with one or more infected persons under three different return-to-school circumstances. Inputs to the infection hazard model are: the inventory of children, teachers and support staff; the prevalence of COVID-19 in the general community including its spatial variation, and the ratio of adult susceptibility to that of children. Three scenarios of inventory are: the counts on 1^st^ June when schools re-opened to Nursery, Reception, Year 1 and Year 6 children, when approximately one-third of eligible children attended; a scenario assuming a full return of eligible children in those cohorts; and a return of all primary age children, scheduled for September. With a national average prevalence, we find that for the first scenario between 178 and 924 schools out of 16,769 in total (i.e. about 1% and 5.5% respectively) may have infected individuals present, expressed as a 90% credible interval. For the second scenario, the range is between 336 (2%) and 1873 (11%) schools with one (or more) infected persons, while for the third scenario the range is 661 (4%) to 3310 (20%) schools, assuming that the prevalence is the same as it was on 5^th^ June. The range decreases to between 381 (2%) and 900 (5%) schools with an infected person if prevalence is one-quarter that of 5^th^ June, and increases to between 2131 (13%) and 9743 (58%) schools for the situation where prevalence increases to 4 times the 5^th^ June level. Net prevalence of COVID-19 in schools is reduced relative to the general community because of the lower susceptibility of primary age children to infection. When regional variations in prevalence and school size distribution are taken into account there is a slight decrease in number of infected schools, but the uncertainty on these projected numbers increases markedly. The probability of having an infected school in a community is proportional to the local prevalence and school size. Analysis of a scenario equivalent to a full return to school with an average national prevalence of 1 in 1700 and spatial prevalence variations, estimated from data for late June, indicates 82% of infected schools would be located in areas where prevalence exceeds the national average. The probability of having multiple infected persons in a school increases markedly in high prevalence areas. Assuming national prevalence characteristic of early June, individual, operational and societal risk will increase if schools reopen fully in September due to both increases in numbers of children and the increased challenges of sustaining mitigation measures. Comparison between incidents in primary schools with positive tests in June and July and our estimates of number of infected schools indicates at least an order of magnitude difference. The much lower number of incidents reflects several factors, including effective reduction in transmission resulting from risk mitigation measure instigated by schools.

## Introduction

On 11 May 2020 Prime Minister Boris Johnson announced that selected primary age children would return to school on 1^st^ June in England. The returning cohort would include Reception, Year 1 and Year 6, noting that there would also be Year 2 to 5 children of frontline workers and those identified as vulnerable who were already being taught in many schools. Nursery age children were also invited to return with some nurseries being part of Primary schools. The devolved administrations decided at the time not to re-open schools. The Government abandoned the proposal for a full return of Primary school children in England before the summer holidays. There is now an expectation that schools will fully re-open in September.

The partial re-opening of Primary schools was widely debated with concerns from some parents, teaching unions and teacher associations about the safety of the children and school staff. There was also concern about the effect on infection rate in the wider community, for example by triggering a second wave. While official guidelines for re-opening schools have been issued by the Department of Education (DfE) and form the policy basis for risk mitigation in schools, individual schools have adopted their own bespoke strategies (Sparks et al. 2020). Some schools re-opened on 1^st^ June, some delayed their re-start while, in other cases, schools did not re-open under advice of local education authorities. Data from DfE indicates on 2^nd^ July that about 88% of state-funded Primary Schools had re-opened to some extent. The community response has been variable and between 1^st^ and 15^th^ June 2020 approximately 35% of eligible children returned. The numbers increased somewhat: 41% (year 1) and 49% (year 6) attended on 2^nd^ July. Between 18^th^ May and 31^st^ July 2020 there have been 247 COVID-19 related incidents in schools of which 116 were tested to be positive test (PHE 2020).

This study is part of a quantitative hazard assessment for schools with respect to COVID-19. The study seeks to enumerate potential infection levels in primary schools, with estimates of attendant uncertainties. The study involves building a stochastic uncertainty model of hazard which includes information on schools and epidemiological parameters that control occurrence and transmission of COVID-19 within schools. Much of the information is derived from national statistics on schools and information about the incidence and prevalence of COVID-19 in England.

This study does not address the implications of opening schools for spread of the infection in the wider community, which are being addressed by other modelling studies. Here we distinguish between societal risk, operational risk and individual risk. The question of how opening schools up affects the national and regional picture of infection is an example of societal risk assessment; for example, by estimating the change in the reproduction number (Panovska-Griffiths et al. 2020). Operational risk concerns a school identifying an infection or the threat of an infection breakout. Mitigating steps might include asking confirmed or suspected COVID-19 persons to self-isolate, classes to be sent home, or even schools to close again, entirely. Individual risk concerns the probability that a child, teacher or other support staff acquires infection as a consequence of the initially unknown presence of an infected person in the school.

Here we have developed a model of infection hazard, which is defined as the probability of one or more infectious person (child, teacher or support staff) being on the premises of a school on any one day. The study uses a fast, stochastic uncertainty modelling tool (UNINET) to construct an uncertainty model for infection hazard levels in primary schools in England. A model of infection hazard is a necessary first step in developing a risk model which needs to include infection transmission. The model is extended to take account of spatial variations in prevalence to characterize infection hazard level in relation to geographical locations. Our model is flexible and dynamic. Thus, in the future it will enable risk to be assessed for different return-to-school scenarios, including secondary schools, for regional or local situations and potentially for individual schools. The approach can also be modified for different kinds of educational operation, such as Universities and Further Education colleges. The model can also be used to estimate sampling power in schools in the context of an infection testing programme.

## Hazard and risk terminology

Different domains of science and society use terms like hazard and risk in different ways and in different contexts. Risk can sometimes be discussed in the context of risk mitigation and safety, whereas in, for example, the financial sector the risk is defined as expected monetary loss. In the health and safety context risk is the chance of harm, whereas in some natural hazard contexts threat is used to signal the potential of harm. Exposure is sometimes used to indicate that an asset or person is in harm’s way. Risk also pertains to different scales and kinds of loss or harm. Risk management benefits from information on the hazard, the exposure, the vulnerability to the hazard and its potential impact. Ultimately, quantitative risk assessment involves calculating the probability of a loss or a harm. Here, we treat COVID-19 infection as a hazard and the presence of infectious persons in schools as a threat that might lead to harm.

## Methodology

Our computational platform is the UNINET LightTwist software (https://lighttwist-software.com/uninet/). UNINET is a standalone uncertainty analysis software with a focus on dependence modelling for high dimensional statistical or probability distributions (Hanea et al. 2018). The program implements and computes continuous non-parametric Bayesian Belief Network (BBN) models as the framework for probabilistic modelling. A BBN is a graphical model, which represents joint distributions in an intuitive and efficient way. It encodes the probability density (or mass) function of a set of variables by specifying a number of conditional independence statements in the form of a directed acyclic graph. The structure of the model is specified by the analyst to incorporate all essential input variables and conditionalizing factors that are needed to quantify the problem and to enumerate output probability distributions. The approach is quite general and allows traceable and defensible results to be generated very efficiently as UNINET uses unique fast Bayesian updating algorithms. One of the other main advantages of UNINET is that it can handle a large number of continuous variables and does not require these to be represented as discretized distributions, as in many BBN programs.

An added benefit is that other factors, elements and complexities can be readily added to the BBN model formulation, for example, by including details of school characteristics or age-dependent transmission rates and, perhaps most pertinently, the BBN calculations can be quickly updated if new or revised data are forthcoming. The model can be applied at different granularities and at a regional or local level. As we have found with volcanic eruptions, this latter capability is invaluable for real-time forecasting applications.

A full list of applications of UNINET to a wide diversity of problems in science, engineering and public health can be found in the reference list on the LightTwist web site. We describe the UNINET platform in Appendix I, explaining the essential calculations, the principles of constructing a simple network as a calculator and provide one explicit example of calculations with an analysis report.

## Data sources

We extracted data on primary schools in England, including their size distributions, from DfE links. Information figures for 2019 can be found at:

https://assets.publishing.service.gov.uk/government/uploads/system/uploads/attachment_data/file/826252/Schools_Pupils_and_their_Characteristics_2019_Accompanying_Tables.xlsx

https://assets.publishing.service.gov.uk/government/uploads/system/uploads/attachment_data/file/812539/Schools_Pupils_and_their_Characteristics_2019_Main_Text.pdf

https://www.gov.uk/government/statistics/schools-pupils-and-their-characteristics-january-2019

https://explore-education-statistics.service.gov.uk/find-statistics/school-pupils-and-their-characteristics

The basic facts about state-funded primary schools in England (2019) are: 16,769 schools; 4,727,090 pupils; and 216,500 teachers.

Data for other support staff to be found at:

https://www.instituteforgovernment.org.uk/publication/performance-tracker-2019/schools

https://assets.publishing.service.gov.uk/government/uploads/system/uploads/attachment_data/file/811622/SWFC_MainText.pdf

https://www.gov.uk/government/statistics/school-workforce-in-england-november-2018

Table 1 shows the numbers of other staff as listed in these sources.

**Table 1.** Data on other support staff in state-funded Nursery and Primary Schools in England (2018 data)

| Staff Categories | Number |
| --- | --- |
| Teaching assistants | 176,679 |
| Administrative staff | 43,771 |
| Technicians | 1,573 |
| Other support staff | 15,240 |
| Auxiliary staff | 44,993 |
| Headcount Third part staff | 26,475 |
| Total | 308,731 |

The Government announcement for re-opening primary schools and nurseries on 1^st^ June stipulated that the returning children would be confined to Reception, Year 1 and Year 6. The pupil database for January 2019 lists numbers of children by age, which does not exactly map onto school years. We used these data to give numbers as follows: 618,700 (Reception), 636,664 (Year 1) and 620,053 (Year 6). A small proportion of Year 2 to 5 children of emergency workers and children from vulnerable environments are present in school. Data for daily national attendance of pupils, teachers, teaching assistants and ancillary staff in schools can be found at:

https://www.gov.uk/government/collections/attendance-in-education-and-early-years-settings-during-the-coronavirus-covid-19-outbreak

For epidemiological data we used a variety of sources. Prevalence data were obtained from infection surveys published by the Office of National Statistics.

https://www.ons.gov.uk/peoplepopulationandcommunity/healthandsocialcare/conditionsanddiseases/bulletins/coronaviruscovid19infectionsurveypilot/latest

ONS state that the number of infections within the community population refers to private residential households, but excludes those in hospitals, care homes or other institutional settings. This measure of prevalence is suitable for our infection hazard model as those presenting at school are not in the excluded group. However, some of these same people will mix with family and friends in the excluded group, suggesting that the national prevalence infection survey data are an underestimate.

Incidence data at UTLA level, to be used as a proxy for regional variations of prevalence, were obtained from Public Health England.

https://www.gov.uk/guidance/coronavirus-covid-19-information-for-the-public

The incidence data were also converted into positive test numbers at UTLA level for the purpose of estimating prevalence and attendant errors in larger scale regions. Population data at UTLA level were obtained from ONS.

## Model Structure

Figure 1 shows the basic Bayesian Belief Network established to estimate infection hazard in schools. Input parameters include prevalence, the susceptibility ratio of children relative to adults in the 20 to 60 year age range, and the inventory of children and staff in Primary Schools. Outputs include the prevalence in schools, the number of schools with one or more infected person on a given day and how the infection is spread between different cohorts of persons (e.g. children and adults).

**Figure 1.**
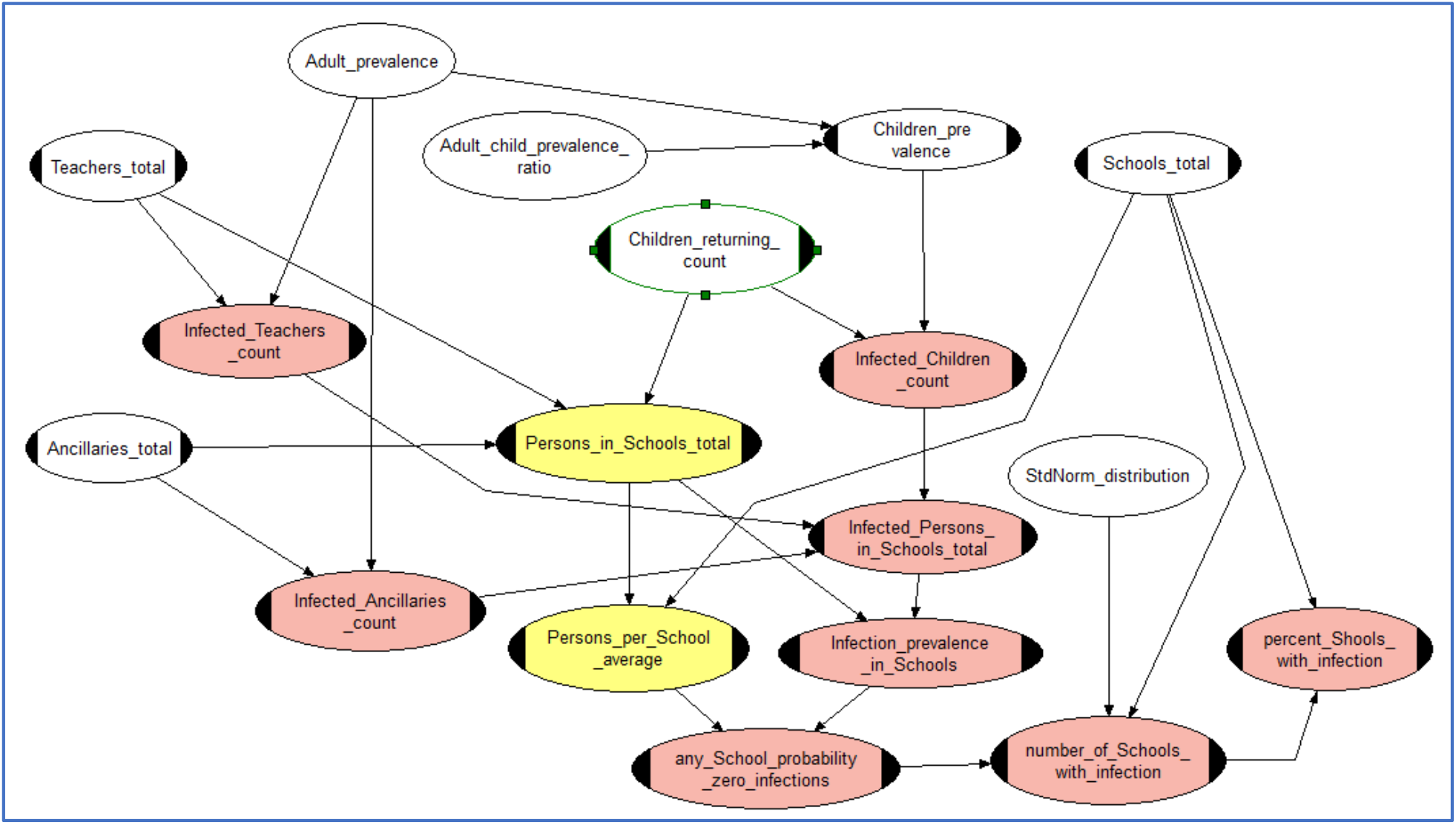
Bayesian Belief Network to calculate schools COVID-10 infection hazard with prevalence represented by a single random variable. In order to account for regional variations of prevalence later BBN modelling introduced a range of random variable values for prevalence in different geographical areas as depicted in Figure 3. Plain white nodes carry input variable uncertainty distributions, white nodes with handles represent fixed (constant) input values. Yellow nodes are intermediate functional (i.e. calculational) nodes. Pink nodes compute the required output variable probability distributions; some these are also intermediate calculation steps, feeding uncertainty distributions into other output nodes.

## Definitions

Having an infectious person in a school poses a threat to individuals, the operation of the school, which in extremis might have to close, and to the wider community. We term a school of one or more infectious individual present as an “infected school.” The harm, potentially caused by this threat, will depend on the context and a full risk assessment would involve quantification of the probability of this harm occurring.

### Scenarios and model assumptions

We adopt here a scenario approach in which some input parameters are either fixed or represented by probability density functions reflecting informational uncertainty. The main assumptions for a given scenario are the number of pupils, teachers, teaching assistants and ancillary support staff present in schools. The scenario parameters are summarized in Table 2 and are now described.

**Table 2.** Inventory of persons present in Primary schools in the model cohorts for the three attendance level Scenarios. All numbers are for England and were extracted from the Government web site and relate to 2020. Scenario I: situation on 1^st^ June 2020 Scenario II assumes attendance of all children in the selected Year groups, adjusted for typical absenteeism (4%). Scenarios IIIa assume a September return of all primary school children in all years, adjusted for typical absentee rates; two sub-scenarios were run for 4 times (Scenario IIIb) and ¼ (Scenario IIIc) the adult prevalence given in the ONS pdf from the 5^th^ June survey.

|  | Scenario I | Scenario I | Scenario II | Scenario III<br>(a, b, c) |
| --- | --- | --- | --- | --- |
|  | Attendance<sup>1</sup> | Numbers<sup>2,3,4,5</sup> | Numbers<sup>3,5</sup> | Numbers<sup>3</sup> |
| Nursery | 17.6% | 55,720 | 297,930 | 297,930 |
| Reception | 29.2% | 194,240 | 626,086 | 626,086 |
| Year 1 | 24.5% | 165,517 | 635,843 | 635,843 |
| Year 2 to 5 |  | 174,938 | 174,938 | 2,327,686 |
| Year 6 | 39.1% | 264,871 | 650,322 | 650,322 |
| Cohort 1 | Rec + Yr1 | 415,471 | 1,559,859 | 1,559,859 |
| Cohort 2 | Yr6 | 439,809 | 825,260 | 2,978,008 |
| Cohort 3 | Teachers +<br>TAs | 235,000±<br>23,500 | 235,000±<br>23,500 | 377,450 |
| Cohort 4 | Support staff | 70,000±7,000 | 70,000±7,000 | 126,720 |

*Scenario I* considers the situation on 1^st^ June 2020, the first day of school return for selected Year groups. Numbers of persons are obtained from the attendance data. The ONS published a probability density function (*pdf*) for adult prevalence on 5^th^ June 2020 (mean value expressed as odds 1 in 1700) and this is used for *Scenario I. Scenario II* assumes attendance of all children in the selected Year groups, adjusted for typical absenteeism (4%). Thus, *Scenario II* represents the case had there been high compliance with the return to school on 1^st^ June.

Looking forward to the full re-opening of schools in September, *Scenario IIIa* assumes the return of all primary school children in all years (adjusted for typical absentee rates). Two additional sub-scenarios are also considered for September: one in which the adult prevalence is 4x greater than that given in the ONS *pdf* from the 5^th^ June survey (*Scenario IIIb*) and another for a drop to ¼ of that prevalence (*Scenario IIIc*). We note here that the prevalence in early August is 1 in 1500, remaining close to the value of 5^th^ June. These sub-Scenarios provide two alternative but plausible ranges of adult prevalence reflecting the future level of, and uncertainty in, prevalence in September. The numbers of persons in each *Scenario* are given in Table 2.

An initial and relatively simple baseline problem for modelling is to estimate the number of schools with at least one infectious pre-symptomatic or asymptomatic individual present. We assume here that once symptoms are recognized steps will be taken to remove the individual from the school and to identify possible contacts. A key issue here is that there is mounting evidence that children are less susceptible to COVID-19 infection than adults. The comprehensive modelling analysis of global data by Davies et al. (2020) led us to adopt a *pdf* for the ratio of adult to children’s susceptibility with a median of 2.3 and 90% credible interval between 1.3 and 3.8 (see appendix 1). This range is within the results of other studies of susceptibility ratio between adults and children (Jing et al 2020; Li et al. 2020; Mizumoto et al. 2002; Zhang et al. 2020).

We divided the persons into four cohorts based on the children’s Year groups and staff roles. While a simplification, the cohorts are expected to have different contact characteristics, e.g. with respect to interactions between children, between children and adults and between adults. The cohorts are as follows: *Cohort 1* are Nursery, Reception and Year 1 children; *Cohort 2* are Years 2-6 children, noting that for *Scenario 1* the Year 2-5 children are those of key workers and from vulnerable environments; *Cohort 3* are classroom teachers and teaching assistants; *Cohort 4* are non-teaching staff such as administrators, cooks, etc, who have more limited contact with children (see Table 1).

In order to develop the basic infection hazard model to take account of regional variations of prevalence, we used incidence data as a proxy for prevalence because there is no directly measured prevalence data at less than national scale. To first order, prevalence can be related to incidence since, at steady state, new infections are balanced by recovered individuals who are no longer infectious. Thus, prevalence can be scaled from incidence data. Table 3 displays the basis for our calibration of regional prevalence from incidence data at an Upper Tier Local Authority (UTLA) granularity.

**Table 3.** Prevalence model derived from incidence data. Data are: weekly incidence (per 100,000) in prescribed bins from PHE data for week starting 25 June 2020 (Column 1); Population within UTLAs having incidence values in the bin range (column 2); population weighting for each bin (Column 3); total number of new cases (positive tests) for one week (Column 4); estimated number of infectious cases in each bin scaled to give bin prevalence of 1 in 1700 (Column 5); sample error based on positive test numbers (Column 6); estimated prevalence as odds (1 in X) in each bin (Column 7); estimated percentage of all infectious persons within each bin (Column 8). Final row are national averages of parameters.

| Weekly incidence | Population | Popn. Weight | New case numbers | Infectious persons | Sample error % | Equivalent Prevalence | % infected persons |
| --- | --- | --- | --- | --- | --- | --- | --- |
| 0.3-1.0 | 3,965,491 | 0.07 | 29 | 180 | 18% | 22,030 | 0.55% |
| 1.0-2.9 | 10,525,879 | 0.19 | 193 | 1,195 | 7% | 8,808 | 3.63% |
| 3.0-9.9 | 23,641,804 | 0.42 | 1,372 | 8,492 | 2.5% | 2,784 | 25.83% |
| 10-29 | 15,128,342 | 0.27 | 2,245 | 13,896 | 1.8% | 1089 | 42.26% |
| 30-100 | 2,280,778 | 0.04 | 975 | 6,035 | 3.0% | 378 | 18.35% |
| >100 | 355,218 | 0.01 | 498 | 3,083 | 4.3% | 115 | 8.68% |
|  | 55,897,512 | 1 | 5312 | 32,881 |  | 1700 | 100.00% |

As with all proxies there are limitations and caveats. Incidence estimated from PCR testing will be underestimated significantly because not all those infected are tested. Further testing is focused more on symptomatic people so many asymptomatic persons may be missing from incidence data. To reconcile incidence and prevalence data from mid-June, for example, it is necessary to invoke a factor of about 6 in the under-reporting of incidence (Table 3). There will inevitably be significant uncertainties in this factor. However, creating an informative proxy model for regional prevalence variation does not require an accurate knowledge of how prevalence and incidence are related because incidence varies by about three orders of magnitude at the UTLA scale. For example, in early July Leicester had a weekly incidence of 140 (per 100,000 people), whereas three other UTLAs reported zero incidence for the same week.

While subject to the above caveats, use of incidence as a proxy enables a simplified model of spatial prevalence distribution to be constructed, scaled by spatial incidence distribution. Incidence and population data from PHE and ONS were processed to create a coarsely binned distribution of prevalence with population weights. Six bins are defined, divided by intervals of weekly incidence per 100,000 persons: 0-0.29; 0.3-0.99; 1-2.9; 3-9.9; 10-29; 30-99; >100 (the latter is only Leicester). Each bin represents an amalgamation of populations data and UTLA incidence rates for the week starting 25^th^ June 2020. Within each bin we estimated incidence from the total number of positive tests and bin population. The error at the 95^th^ confidence level was calculated from the total number of positive tests sampled within each bin. Table 3 shows the data and the caption describes the basis for the model of regional prevalence.

A final embellishment added to the model explored the effects of school size distribution. Our baseline scenarios assumed all primary schools had a mean pupil population of 282 and averaged numbers of classroom and support staff based on the data in Table 1. The size distribution of primary schools in England based on pupil numbers is wide and polymodal (Figure 2).

**Figure 2.**
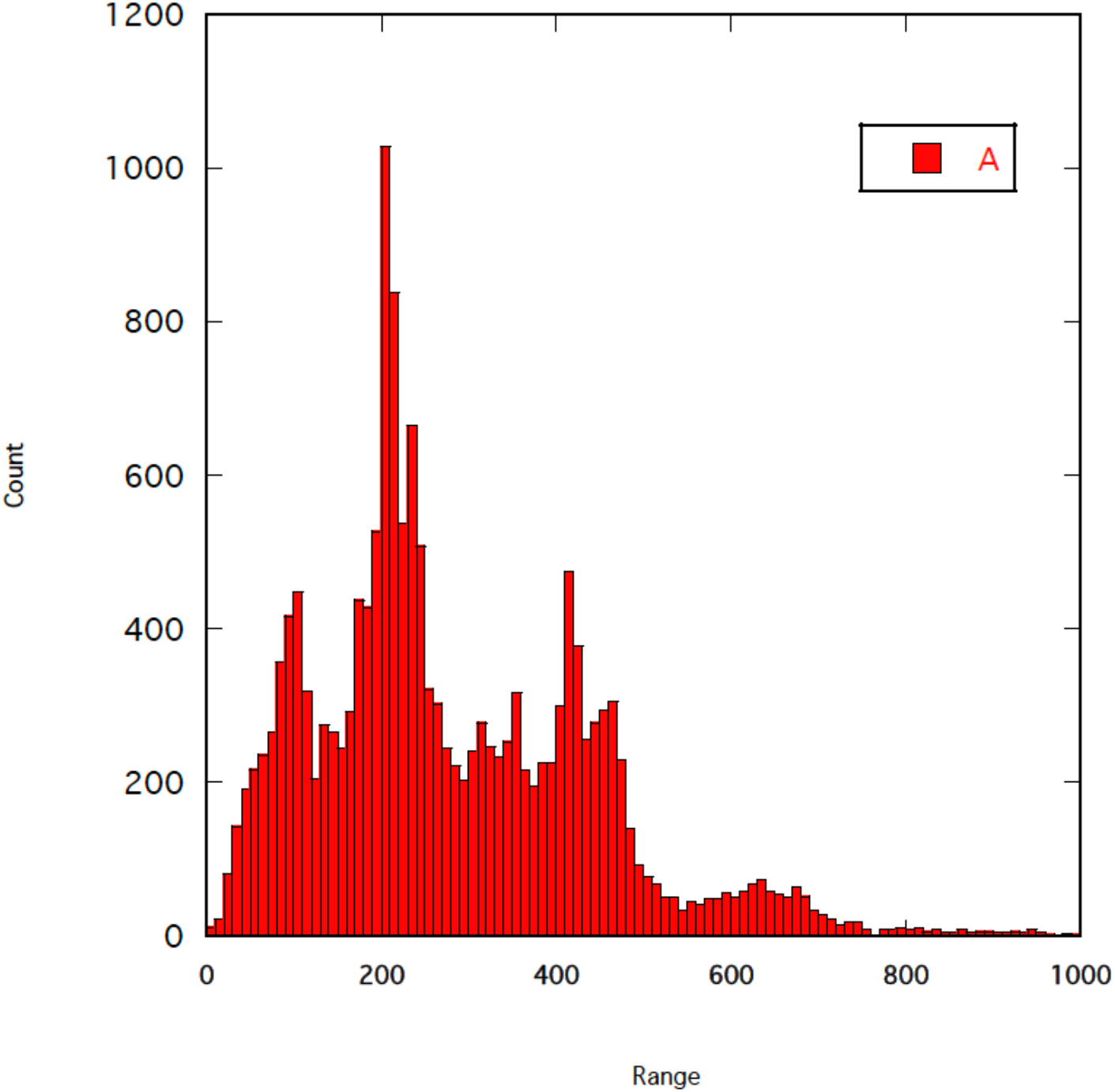
Size distribution of Primary Schools in England based on pupil numbers.

## Results

In the following findings, we report our results as values rounded to an appropriate number of significant figures. Given the large uncertainties present in the results, exact values - as provided automatically by the modelling program - can give a misleading impression of precision and accuracy when output is reported without qualification. For brevity, we discuss some of the results and compare *Scenarios* in terms of median and mean values. However, it is essential that the large uncertainties in the model outputs are fully appreciated.

### Infection hazard levels

We first present results for the distribution of infected schools for the three defined *Scenarios*. Numbers of schools with one or more infected persons are listed in Table 4a and counts of infected children, teachers and support staff, present in the national primary school system, are given in Table 4b. Example model output distributions are shown in Figures 3a-c, illustrating typical uncertainty spreads and some skewness in distribution shapes; long-tailed behavior, as evinced here, could have implications for decision-making. Percentages of infected schools are given. As expected, the relative number of schools with infected persons increases in proportion to the increasing number of returning children and teachers.

**Table 4a.** Infection hazard results: number of state primary schools with one or more infected persons present (with percentage of all schools in parentheses).

|  | Mean | Std dev | 5% quantile | 50% quantile | 95% quantile |
| --- | --- | --- | --- | --- | --- |
| *Scenario I* | 460 (2.7%) | 244 | 178 (1.1%) | 406 (2.4%) | 924 (5.5%) |
| *Scenario II* | 911 (5.4%) | 503 | 336 (2.0%) | 798 (4.8%) | 1873 (11.1%) |
| *Scenario IIIa* | 1635 (9.8%) | 732 | 612 (3.6%) | 1444 (8.6%) | 3310 (19.8%) |
| *Scenario IIIb* | 5255 (31%) | 2350 | 2131(12.7%) | 4863 (29.1%) | 9743 (58.3%) |
| *Scenario IIIc* | 437 (2.6%) | 243 | 162 (0.9%) | 381 (2.3%) | 900 (5.3%) |

**Table 4b.** Estimated number of infected persons in primary schools in England, by return/attendance Scenarios

|  | Number | Std dev | 5% quantile | 50% quantile | 95% quantile |
| --- | --- | --- | --- | --- | --- |
| ***Scenario I*** |  |  |  |  |  |
| Children | 267 | 166 | 88 | 227 | 582 |
| Teachers & TAs | 155 | 78 | 64 | 139 | 302 |
| Ancillary staff | 46 | 23 | 19 | 41 | 90 |
| ***Scenario II<sup>1</sup>*** |  |  |  |  |  |
| Children | 745 | 464 | 246 | 632 | 1624 |
| ***Scenario IIIa*** |  |  |  |  |  |
| Children | 1417 | 883 | 468 | 1203 | 3090 |
| Teachers & TAs | 249 | 125 | 102 | 223 | 487 |
| Ancillary staff | 83 | 42 | 34 | 75 | 160 |
| ***Scenario IIIb*** |  |  |  |  |  |
| Children | 5458 | 3558 | 1713 | 4572 | 12188 |
| Teachers & TAs | 960 | 511 | 372 | 847 | 193 |
| Ancillary staff | 322 | 171 | 125 | 284 | 648 |
| ***Scenario IIIc*** |  |  |  |  |  |
| Children | 360 | 219 | 121 | 307 | 776 |
| Teachers & TAs | 63 | 31 | 27 | 57 | 121 |
| Ancillary staff | 21 | 10 | 9 | 19 | 41 |

**Figure 3.**
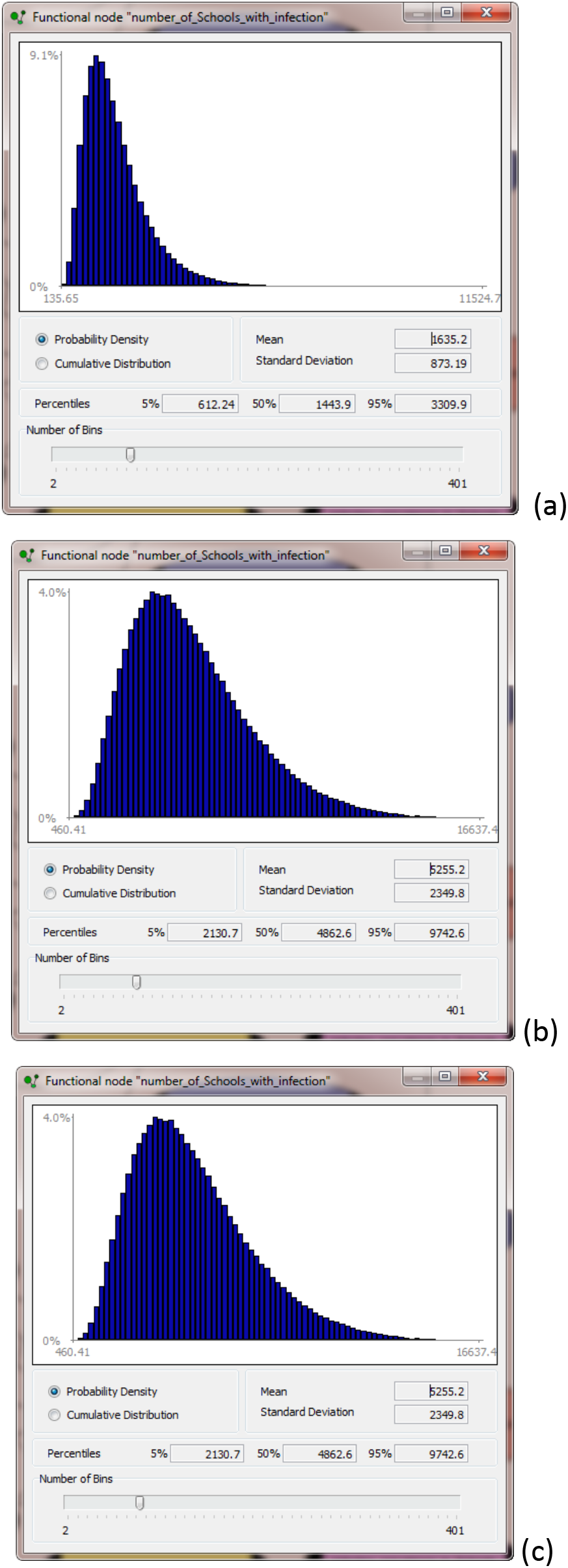
Scenario IIIa: probability density functions for number of Schools with infection, using the model of Figure 1 (results should be read to nearest whole number - see Table 3a) (a) Scenario IIIa; (b) Scenario IIIb; (c) Scenario IIIc.

The uncertainties are large and so only allow rather general inferences. Assuming a fixed prevalence (5^th^ June 2020 ONS data) under current conditions (*Scenario* I), a few hundred schools are likely to be infected, while with full return (*Scenario IIIa*) the proportion increases by a factor of slightly over 3.6x, in terms of mean numbers of schools involved. The increase in number of persons between *Scenario I* and *Scenario III* (a factor of 5.6x) is greater than the projected increase in the number of infected persons. This result is a consequence of the lower susceptibility of children: the bulk of the increase in persons in school, between *Scenario I* and *Scenario III*, is dominated by children, since about 65% of adult staff have already returned.

Prevalence in the community is a major factor in infection hazard. The large uncertainties in the ONS national prevalence infection surveys make it difficult to infer trends. Prevalence has fluctuated modestly over the last several weeks, declining through June and early July but increasing during late July and early August at the time of writing. Thus, June 5^th^ data appear representative and are similar to early August. The effects of prevalence are clear from comparing *Scenarios IIIa, IIIb* and *IIIc* (Table 4b). If continued public adherence to guidance is maintained to September and prevalence drops to ¼ of current levels then, in terms of mean values, approximately 2.5% of schools will have infected persons, compared to 10% at current levels. However, a second wave, causing, say, a factor of 4 increase in prevalence, will increase this estimate to about 30%. We stress that uncertainties are large so there is a 1 in 20 chance that the percentages could be higher than the 95^th^ percentile estimate reported in Table 4a (i.e. 58%). The prevalence in schools is lower than in the general community because children dominate the school population. If the general community prevalence has a median value of 1 in 1700 then the median value of prevalence in schools is estimated as 1 in 2800 for *Scenario I* and 1 in 3300 for *Scenario IIIa*. A major caveat, addressed below, is that prevalence may vary considerably from region to region, or even from locality to locality, and there may be very localised infection hot spots. This is one limitation to calculating - and relying on - averaged or overall national values.

Infection hazard for a particular school is a function, *inter alia*, of size. The average Primary school in England hosts about 280 children. The percentages in Table 4a can be applied to a school of this average size to give the probability that such a school will have an infected person on any one day. The mean odds are 1 in 40 for *Scenario I* and 1 in 10 for *Scenario IIIa*. Using children count as an approximate proxy for school size, the mean odds of there being an infected person present in a small rural school with 60 children would be about 1 in 190 while, for a large urban primary school of 700 children, the corresponding probability would be 1 in 16.

The number of infected persons in Table 4b add up to a slightly larger count (by a few) than the number of schools in Table 4a. This difference is due to a small number of schools which will have two or more infected persons present on the same day. Conditional on at least one infected person being present, the probability of having two or more cases in one school of average size is approximately 0.005 (i.e. 1-in-200), while for three or more it is about 0.00015 (1-in-6667). However, these probabilities only apply to infected persons drawn at random from the wider community, assuming average national prevalence; the risk of having multiple infected persons in a school is developed further, below, in relation to spatial prevalence variations. These estimates do not take account of secondary infections occurring within the school. Our analysis also assumes that all individuals (pupils, teachers and other staff) are independent ‘samples’ in population statistics terms, and therefore their probabilities of infection are uncorrelated. If there were two or more siblings in one school, say, then the joint probability of them being infected together is increased relative to two unrelated or otherwise unconnected members of a population (see Appendix 2).

Infection hazard does not equate directly to the extent that infection might spread, but we expect that risk of an outbreak will be proportional to infection hazard. Between 18^th^ May and 31^st^ July 2020 there have been 247 COVID-19 related incidents in schools of which 116 were tested to be positive test (PHE 2020). Details about the circumstances for those that tested positive are not accessible, but the data provide a lower bound on the number of schools with infectious persons present. There may be a significant number of asymptomatic persons present and it also does not follow that having an infectious person in a school will lead to an incident leading to a test, especially in the context of stringent risk mitigation measures being applied in primary schools. The number of schools estimated to contain an infectious person on a given day is thus much higher than the occurrences of reported cases with positive tests.

There are many reasons why infection may not be passed on, resulting in a much reduced frequency of outbreaks and hence lower disease incidence. The infectiousness of the particular individual may be low and they may start feeling ill outside school hours. Infectiousness is thought to be at a maximum just before feeling ill (He et al. 2020; Ashcroft et al. 2020) and this heightened state might be reached also outside school hours (e.g. weekend). In contrast, asymptomatic carriers may not be recognized at all. The risk mitigation measures to reduce contacts between people, ensure hygiene and isolate persons who display possible COVID-19 symptoms (Sparks et al. 2020) indicate they are effective in reducing opportunities for infection spread.

It is tempting to scale up from the schools’ actual breakout data during *Scenario I* to project potential *Scenario III* incidents in a linear manner, using the relative exposed population ratio. However, it seems more likely that the scaling will not be linear. In particular, risk mitigation might be less strict and harder to maintain when many more children are back in school.

### Spatial variations in prevalence

The results in Tables 4a and 4b, and discussed above, assume a constant prevalence across the UK, but this is clearly not the case with localized hotspots, such as Leicester, Blackburn and Bradford in July. Regional variations of prevalence in early July suggest a variation at the UTLA (ONS upper tier local authority) scale of at least 350x. We have explored as a simple illustration how this variation might affect the results. In the BBN, six half-log incidence bins with sampling uncertainties are introduced, with separate contiguous weekly incidence rates, and these are converted to equivalent prevalence distributions using a generic scaling factor (Table 3); the prevalence values are the inverse of the odds of an infected person in a population. For each bin we calculated from ONS data the total number of persons that tested positive in each bin to estimate a sampling error.

We have modelled regional prevalence variation for a case equivalent to *Scenario IIIa* with a uniform prevalence. Table 5a shows prevalence rates partitioned into six regions (Bins) with the quantiles of the distributions ascribed to each Bin. Also shown are the relative population weightings for each Bin. These BBN node histogram distributions are shown in Figure 4, with means and standard deviations in the lower panels.

**Table 5a.** Model spatial distribution of prevalence dividing England into six contiguous half-log prevalence regions, with weightings reflecting the relative population sizes of these regions. The net mean prevalence in the BBN, in odds terms, is slightly higher than the corresponding overall mean from the tabulated data (1-in-1631 -v- 1-in-1700). This is likely due to slight misfits when converting Bin raw data into separate statistical distributions.

| Prevalence | Data mean | BBN mean | 5th %ile | 50th %ile | 95th %ile | Population weighting |
| --- | --- | --- | --- | --- | --- | --- |
| **Bin 1** | 3.60E-05 | 3.48E-05 | 0 | 4.20E-05 | 6.12E-05 | 0.07 |
| **Bin 2** | 1.20E-04 | 1.17E-04 | 6.95E-05 | 1.12E-04 | 1.81E-04 | 0.19 |
| **Bin 3** | 3.42E-04 | 3.33E-04 | 1.94E-04 | 3.18E-04 | 5.24E-04 | 0.42 |
| **Bin 4** | 9.54E-04 | 9.48E-04 | 5.71E-04 | 9.10E-04 | 1.45E-03 | 0.27 |
| **Bin 5** | 2.59E-03 | 2.56E-03 | 1.59E-03 | 2.47E-03 | 3.85E-03 | 0.04 |
| **Bin 6** | 8.41E-03 | 8.41E-03 | 5.46E-03 | 8.41E-03 | 1.14E-02 | 0.01 |
| Overall means | 5.88E-04 | 6.08E-04 |  |  |  |  |
| **Odds 1-in-** | **1700** | **1645** |  |  |  |  |

**Figure 4.**
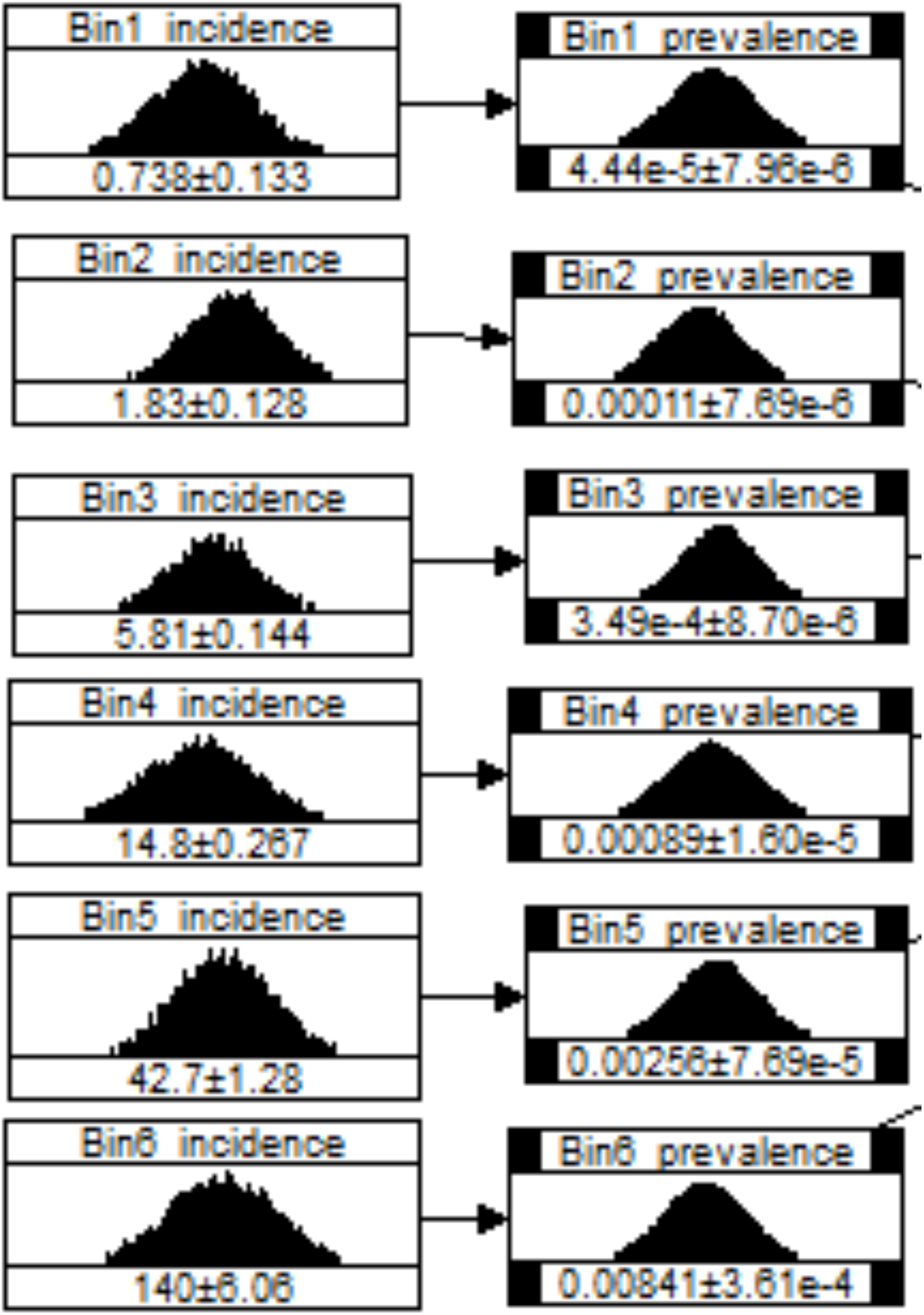
Node distributions representing regional variations in incidence, converted prevalence and associated uncertainties used in the BBN infection hazard model (see Table 5a). These nodes replace the single distribution node “Adult-prevalence” in the basic BBN shown in Figure 1, above.

In terms of schools’ infection hazard results, Table 5b shows the statistical characteristics of the estimated number of state primary schools with one or more infected persons for the *Scenario IIIa* base model versus the multi-prevalence model, outlined above (see also Figure 4); the equivalent percentages of all schools are shown in parentheses. The prevalence model with spatial variation projects slightly reduced number of infected schools compared with the uniform prevalence model. Estimated number of infected persons in primary schools, by pupil, teacher or ancillary staff, for *Scenario IIIa* base model versus multi-prevalence model are shown in Table 5c. In terms of mean values, the numbers are almost identical. However, the variances on the number of schools and infected numbers of persons in the spatial model are increased relative to the base model with more marked skewness reflected in the lower median values compared to the uniform prevalence model.

**Table 5b.** Infection hazard results: number of state primary schools with one or more infected persons present (with percentage of all schools in parentheses): Scenario IIIa base model versus multi-prevalence model versus multi-prevalence model including school size distribution (Figure 2).

|  | Mean | Std dev | 5% quantile | 50% quantile | 95% quantile |
| --- | --- | --- | --- | --- | --- |
| **Scenario IIIa** | 1635 (9.8%) | 732 | 612 (3.6%) | 1444 (8.6%) | 3310 (19.8%) |
| **Spatial prevalence** | 1458 (8.7%) | 1873 | 122 (0.7%) | 888 (5.3%) | 4344 (25.9%) |
| **School size distribution** | 1405 (8.4%) | 1910 | 59 (0.35%) | 754 (4.5%) | 4910 (29.3%) |

**Table 5c.** Estimated number of infected persons in primary schools: Scenario IIIa base model versus multi-prevalence model (Figure 3).

|  | Number | Std dev | 5% quantile | 50% quantile | 95% quantile |
| --- | --- | --- | --- | --- | --- |
| **Scenario IIIa** |  |  |  |  |  |
| Children | 1417 | 883 | 468 | 1203 | 3090 |
| Teachers & TAs | 249 | 125 | 102 | 223 | 487 |
| Ancillary staff | 83 | 42 | 34 | 75 | 160 |
| **Spatial prevalence** |  |  |  |  |  |
| Children | 1415 | 2699 | 95 | 726 | 4167 |
| Teachers & TAs | 248 | 443 | 19 | 136 | 652 |
| Ancillary staff | 83 | 149 | 6 | 46 | 219 |

To model the distribution of infected schools in areas of different prevalence, we individually modelled each bin with school numbers proportioned to bin population. Table 6 shows the results comparing the bins by number of infected schools per 100,000 people. One major and unsurprising consequence of spatial variation of prevalence is that the number schools with infectious individuals is strictly proportional to prevalence. The corollary of this result is a lower mean number of schools with infection (and lower numbers of infectious pupils, teachers and ancillaries) in areas of lower prevalence. Figure 5 displays the results as percent of infected schools that exceed a given prevalence value. For this illustrative scenario, 82% of infected schools are in areas of above average prevalence.

**Table 6.**
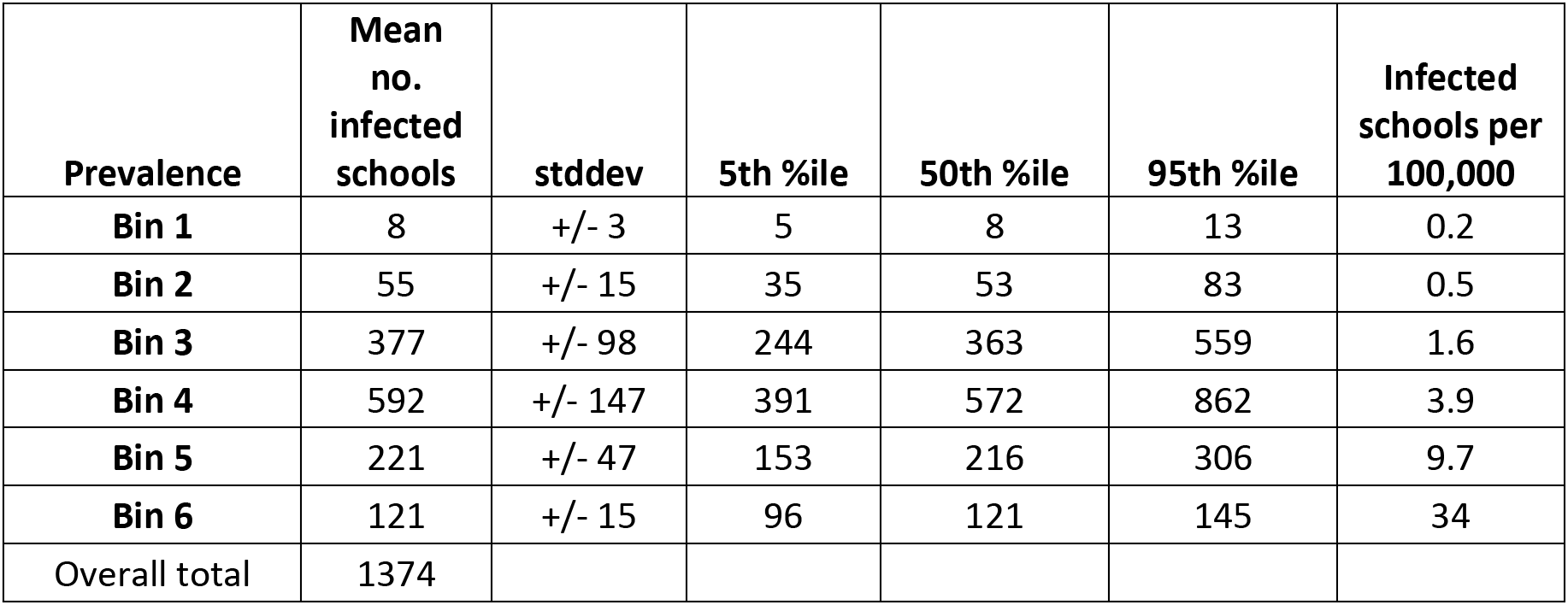
Distributions for estimated average number of schools with one or more infected person as a function of spatial prevalence levels; calculations are based on the assumption that the total numbers of schools per prevalence bin are proportional to the corresponding population size. Different school size profiles within prevalence bins are not accounted for here, so the tabulated numbers of infected schools relate to an average. sized school.

| Prevalence | Mean no. infected schools | stddev | 5th %ile | 50th %ile | 95th %ile | Infected schools per 100,000 |
| --- | --- | --- | --- | --- | --- | --- |
| Bin 1 | 8 | +/- 3 | 5 | 8 | 13 | 0.2 |
| Bin 2 | 55 | +/- 15 | 35 | 53 | 83 | 0.5 |
| Bin 3 | 377 | +/- 98 | 244 | 363 | 559 | 1.6 |
| Bin 4 | 592 | +/- 147 | 391 | 572 | 862 | 3.9 |
| Bin 5 | 221 | +/- 47 | 153 | 216 | 306 | 9.7 |
| Bin 6 | 121 | +/- 15 | 96 | 121 | 145 | 34 |
| Overall total | 1374 |  |  |  |  |  |

**Figure 5.**
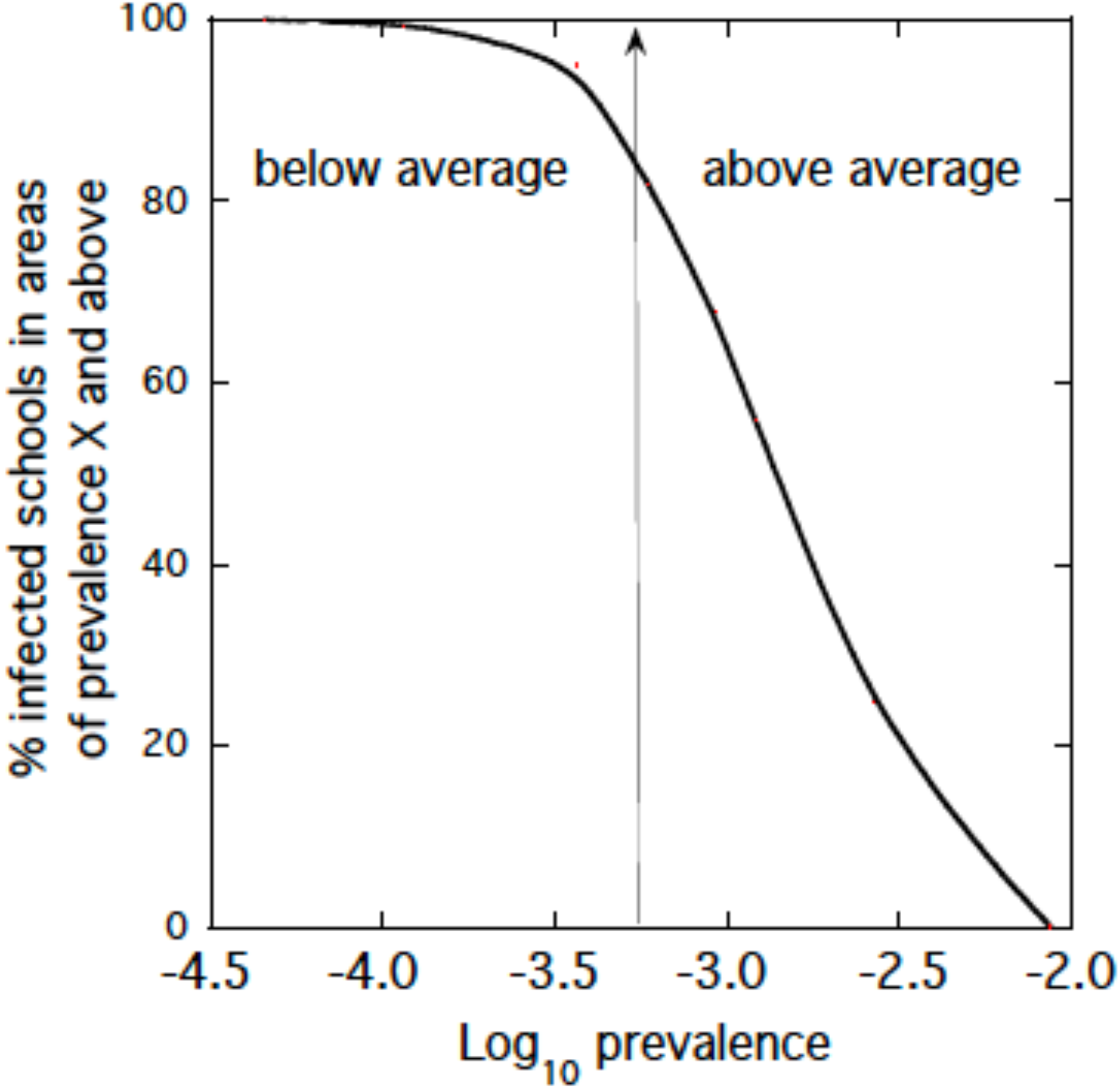
Survivor function plot showing percentage of infected schools in areas of prevalence X and above. The percentage of infected schools that occur at or above a fixed value of prevalence can be read off the curve. The model is *Scenario IIIa* with regional prevalence variation. The central vertical line is the national average prevalence assumed in the model. 82% of infected schools are located in areas with prevalence above the national average.

One of the consequences of high prevalence in an area is to concentrate, in that area, the number of schools that have two more infectious persons on the same day. An approximate measure of the number of schools that have multiple infectious persons is the difference between the number of schools with one or more infected persons and the corresponding infected person numbers: the excess infectious cases indicate some schools in a high prevalence area can expect to have two or more cases at any one time. For the uniform model (*Scenario IIIa*), the difference between number of schools with an infected person and number of infected persons is 114, but for the spatial prevalence model the difference is 288. This result is significant in that there will be almost a factor of 3 increase in risk of an outbreak in those schools with multiple infected persons.

### Effects of school size and multiple infections

Schools vary significantly in size (Figure 2). We expect that the probability of encountering one or more infected persons in a school will depend on size. We therefore ran a model which sampled from the national distribution of school sizes, rather than assuming a mean school size. The model chosen was *Scenario IIIa* with spatial prevalence variation. The results are shown in Table 5b. Including variation in school size reduces the number of infected schools from 1458 to 1405 while the uncertainty range increases slightly. The decrease is because of the higher probability of an infectious person being encountered in a larger school. Since there is no change in the overall number of infectious persons, the implication of a smaller mean number of schools with infection is that there will more cases of multiple infections in some schools.

The discussion has so far focused on estimating the probabilities of encountering one or more infected persons in school. Because there could be implications for transmissibility risk levels, our analysis is extended to enumerate probabilities for 2, 3, 4 … N infected persons per school. For the model including the school size distribution and regional prevalence we find mean proportions as follows for *Scenario IIIa*:

> N = 0 Prob = 0.916
>
> N = 1 Prob = 0.072
>
> N = 2 Prob = 0.009
>
> N = 3 Prob = 0.002
>
> N = 4+ Prob = 0.001

By way of illustration, we contrast a small school (50 pupils; 10 teachers; 10 other staff), in an area with the second lowest spatial mean prevalence, with a large school (900 pupils; 100 teachers; 100 other staff) in the second highest prevalence area (see Figure 3). The probability of one infected person (pupil or adult) in the small school is 4.8×10^-3^, for two or more the probability is about 1.1×10^-5^. In contrast, for the large school the probabilities of N infected persons are:

> N = 0 Prob = 0.26
>
> N = 1 Prob = 0.35
>
> N = 2 Prob = 0.24
>
> N = 3 Prob = 0.11
>
> N = 4 Prob = 0.04
>
> N = 5+ Prob = 0.01

For a very big primary school in a high prevalence area, there is a 40% chance of having 2 or more infected persons and about a 1% chance of encountering five or more infected persons in the school at any one time.

### Comparison with observations

Our model for *Scenario Ia* gives a mean of 460 (90% credible range 178 .. 924) infected schools on any one day. Taking account of regional prevalence variations and school size distributions this expected number would be somewhat reduced by about 15% to 390 (see Table 5b). With time the numbers of schools in which an infected person has been present will slowly grow so an implication is that the number of schools will be much higher after a few weeks. A quite complicated calculation would be needed to infer what numbers of primary schools could have been infected in June and July. It would need to consider both temporal and spatial variations of prevalence and variations of how long an individual remains infectious. A very approximate simplified calculation, however, gives a sense of the number. If infectiousness typically lasts a week (He et al. 2020) then for fixed prevalence and a daily rate of new infected schools of 55 per day, the total number of schools that had been infected would be about 1925.

Between 18^th^ May and 31^st^ July 2020, 247 COVID-19 related incidents were reported in schools, of which 116 were tested to be positive test (PHE 2020). Even allowing for the large uncertainties, these numbers are below our estimate of the number of infected schools. However, it seems likely that many infections were not detected because the infected persons were asymptomatic or developed unreported symptoms outside school hours. The low number could also reflect incomplete testing coverage. Further, the probability of transmission could be significantly lower than 1. We have shown elsewhere (Sparks et al, 2020) that close contacts between persons in schools were reduced in Primary schools by between 45% and 75%, compared to normal Pre-COVID times, while other risk mitigation measure will have reduced transmission significantly.

## Use of findings for informing policy decisions

The main policy implications of our preliminary infection hazard results are that in England a few hundred primary schools were expected to have infectious individuals within them in early June, just after re-opening. With a partial return to school, social distancing measures are relatively easy to comply with. On this basis, for a full return to school in September, similar numbers of schools are expected to have infectious persons present on one day, but only provided that prevalence falls to about ¼ of the 5^th^ June level; on the other hand, numbers can be expected to more than triple if prevalence stays at current levels.

If a national scale second wave comes with, for example, community prevalence increasing by a factor of 4 compared to 5^th^ June, several thousand schools may have infectious persons present. This despite the fact the net prevalence in schools is likely to be much lower than in the general community because of the high proportion of persons in schools being children with lower infection susceptibilities. Infection hazard is proportional to school size but also depends on the relative proportion of children and adults. Social distancing measures will be much harder to implement with a full return to school.

A model which explores the effects of spatial prevalence variations finds that schools with infected persons are preferentially concentrated in high prevalence communities. There is an overall slight decrease in the number of infected schools nationally. However, in high prevalence areas there is an approximately three-fold increase in likelihood of some schools having more than one infectious person in attendance at any one time, even without considering the effect of siblings and transmission leading to secondary infections. Infections will tend to be concentrated in larger schools.

In the *Scenario IIIa* model with regional prevalence variations, the number of infected schools per 100,000 people varies by a factor of over 350 between the lowest and highest prevalence areas in England at the UTLA scale. This large variation suggests a strong policy focus on mitigating risk in high prevalence areas. In our illustrative model, 82% of infected schools are located in areas with prevalence above the national average. One option for government or for a local authority is to consider stricter risk management regimes in schools where and when local incidence rates exceed a threshold. To set such a threshold on a reasoned and defensible basis, requires a formal quantitative risk assessment, with uncertainties properly incorporated.

Our results are also consistent with the effectiveness of risk mitigation measures taken in schools to reduce transmission. The number of incidents in schools with positive tests are less than the number of expected schools with infectious persons present by at least an order of magnitude. Unfortunately, interpretation of the difference is confounded by other factors (e.g. completeness of testing and role of asymptomatics). Nonetheless the reduction of contacts within primary schools of between 45 and 75% (Sparks et al. (2020) and instigation of other safety measure (e.g. handwashing and fastidious cleaning) supports risk mitigation as a significant factor.

## Further work

One factor not fully explored here is the size distribution of primary schools. We have assumed that the national size distribution applies everywhere, but this size distribution is likely to vary at the UTLA scale across the country, depending on the character and make-up of the local community Thus, our results can be anticipated to change somewhat when repeated at greater granularity at Lower tier or school catchment scale, for example. A city like Leicester (a single UTLA) could be modelled at much finer local scale to understand which individual schools within that conurbation will have higher COVID-19 infection hazard. Our model captures the first order picture but does not have the necessary spatial resolution to understand infection hazard at local levels. Postcode-based incidence and prevalence data would allow modelling to address risks at an individual school’s catchment scale.

For incidence parameter uncertainty characterization, we adopted the assumption of lognormal distributions for the different partitions of the UTLA geographic-referenced data. As always, there are a number of reasons for settling on one preferred distribution type or another; in many disciplines, when treating positive real variable data, a lognormal process is regarded as the statistical realization of the multiplicative product of several factors, which is likely to be true with incidence/prevalence data. The lognormal has the advantage of common statistical usage, and understanding, hence we adopted here for our initial ‘scoping’ analyses. Other distributions (e.g. Weibull; gamma) may represent more appropriate functional forms for these particular data when aspects like goodness-of-fit are considered. We intend to explore alternatives objectively in follow-on work, paying attention to upper tail fitting because, in a safety-critical risk assessment, it is there that low probability, but potentially disastrous outcomes may reside. The likelihood of occurrence of such threats warrants enumeration as reliably as possible, in the face of many intrinsic uncertainties.

Our BBN model, as now set up, can also be configured straightforwardly to estimate infection risk in schools of specific sizes (i.e. small, medium or large) and for individual schools, of given size, in areas with known, specific prevalence rates. The model can be operationalized at national and local levels to give near real-time infection hazard informed by weekly changes of incidence and prevalence. The model is thus suitable for decision support with regard to management strategies at school, city, local authority and national scales, and for informing other research programmes, e.g. infection testing in schools.

This interim report only presents the results of an initial first stage analysis. The school infection rate estimates presented above do not include the possibility of individual schools having sibling pupils who may be simultaneously infectious; these would need to be modelled as nonindependent cases, i.e. correlated samples, within a school population. In similar vein, there can be a number of schools with twins or triplets in the same class; this, too, would affect potential infection and onward transmission rates. An initial scoping exercise, however, suggests that co-sibling infection rates are low and, as the proportion of a school pupil population that is represented by siblings is also small, the influence of children living at the same home address is likely to be marginal for school population infection rates.

The current model is not a direct measure of outbreak risk in schools, although we surmise the infection hazard is strongly correlated with outbreak risk. We have noted that the central (mean) model estimates the daily presence of a few hundred infected primary schools in England during June. Reported incidents with associated positive tests amounted to 116. While hypothesizing that infections are likely to have been undetected or under-reported and while transmission has been strongly suppressed by risk mitigation measures applied in the schools, much more work needs to be done to understand the differences. An adjunct transmission model will need to be added to our infection hazard to characterize the probability of detected infections outbreak, given the presence of one or more infectious individuals in a school.

## Data Availability

Data are all from publicly available sources and are cited within manuscript

## Acknowledgements

The study was part of the RAMP initiative of the Royal Society. We thank Leon Danon and Jonty Rougier for commenting on an earlier version of this report.

## Appendix 1. Stochastic Modelling and Uncertainty Analysis with UNINET

In the present study, we use UNINET in its basic stochastic uncertainty modelling mode, i.e. to perform Monte Carlo re-sampling from random variables described by statistical distributions or by uncertainties derived from expert judgement elicitation. (In this application, the program is not used for Bayesian inference, although the model is represented by a directed acyclic graph very similar to a Bayesian Belief Network, a BBN).

In the following simple illustrative model (Figure A1), there are two random variables, one a Lognormal distribution, the other a Normal distribution, each with definable parameters. These ‘parent’ nodes are connected to a third ‘child’ node by influence arcs, denoting the child node has dependencies on both parents. The child node is a functional or calculational node, that is, it contains a formula for solving its relationship to the parents, in this case, the simple product LogNormal * Normal.

The result of performing multiple re-sampling of the two parent node distributions and calculating the product of these samples in the third node computes the full statistical distribution of the product, given the uncertainties associated with the parent nodes (in this model, 100,000 samples were used). UNINET can show graphically all node distribution shapes, with their means and standard deviations displayed, as below, and can report more detailed properties of the distributions, as in the lower right panel; the program can also export the individual samples collected from the Monte Carlo re-sampling for post-processing in other forms of analysis.

**Figure A1.**
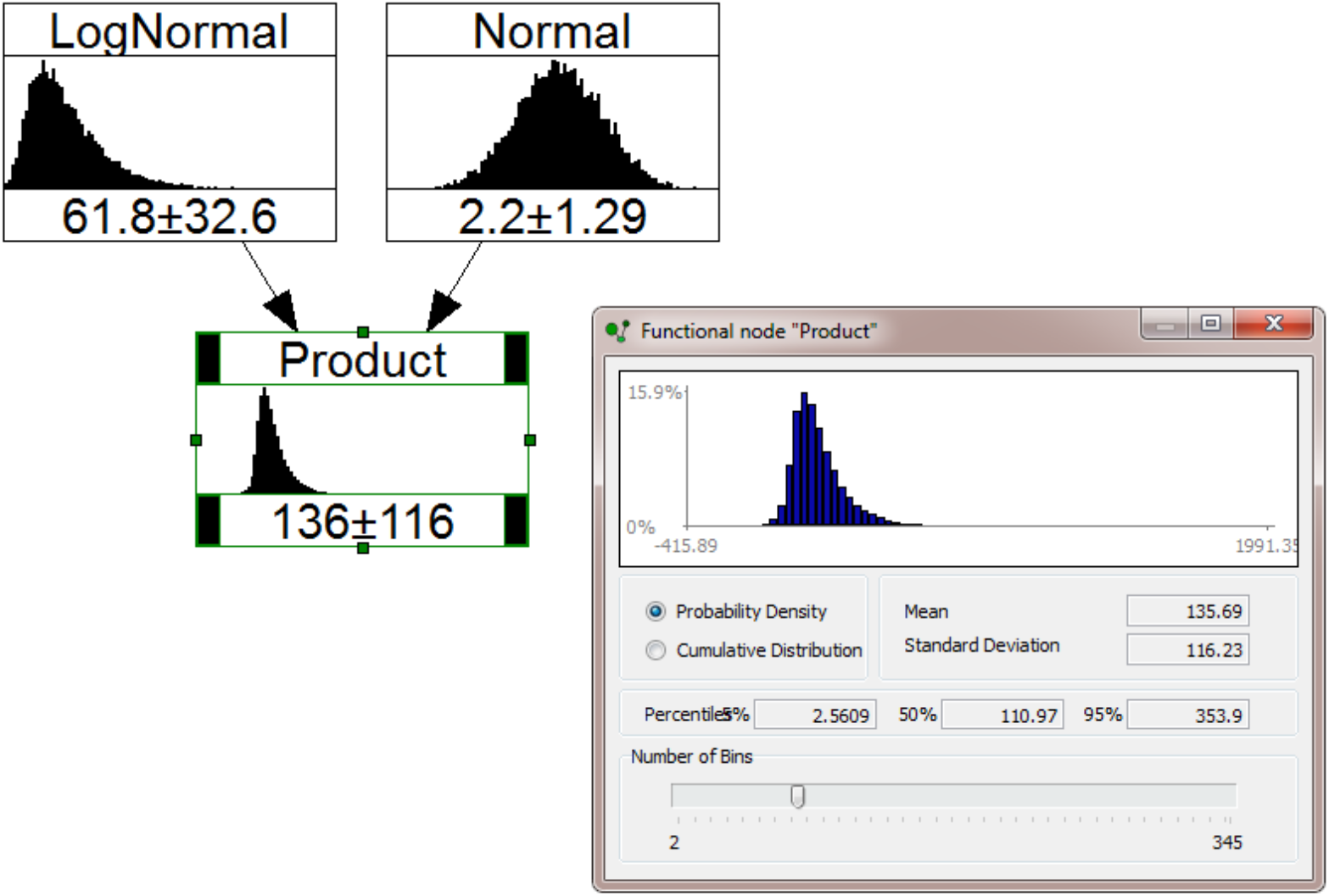
Simple UNINET model showing how variable uncertainty distributions can be represented by probability nodes (e.g. LogNormal, Normal) and calculations can be made with functional nodes containing equations or formulae (e.g. node Product). For a set number of sampling iterations, the Product node computes the product of LogNormal * Normal and creates a new distribution reporting its mean and quantiles.

The UNINET program includes implementations of several common forms of statistical distribution, which can be uniquely parameterised, and has a full range of numerical operators for performing stochastic calculations via functional node equations defined by the analyst. In addition, the program has options for computing results when input variables are conditionalized on specific point values or ranges of values. With these powerful capabilities, UNINET provides an efficient, easily developed modelling tool for conducting sophisticated exploratory uncertainty analysis in problems of high dimensionality.

The calculations being performed within the UNINET platform in this study would be exceedingly simple if there were no uncertainty in the pertinent epidemiological parameters, no spatial variations in prevalence and if all schools were exactly the same size with the same proportions of children, teachers and other support staff. In addition, we need the total number of schools (N_s_), children (N_c_) and adults (N_a_) in the school system. Such a simple but unrealistic case, however, does allow the essence of the calculations to be described. For schools of identical characteristics the number of infectious persons present in schools would be proportional to prevalence, p, and would depend on the susceptibility ratio of the adults relative to children, S_r_. For simplicity the susceptibility of an adult is taken as 1. The number of infectious children, I_c_, and infectious adults, I_a_, can then be written as:

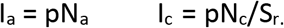

Even in this simplest of case the number of schools with an infectious person (I_s_) would not necessarily equal the sum of I_a_ and I_c_ because of the possibility that, by random chance, a few schools might have 2 or more infectious people. Thus, we could expect I_s_ ≤ I_a_ + I_c_. For very small p <<1, I_s_ ~ I_a_ + I_c_. However, even here a computational model would need to be developed to randomly assign infectious persons among the Ns schools and repeated calculations would result in uncertainty for I_s_.

However, the key epidemiological parameters have large uncertainties, while school sizes and relative proportions of children and adults vary. Therefore, what is required is a high dimensional computation enabled by the UNINET platform in which the uncertainty distributions are sampled a very large numbers of times in order to make sure that tails of the probability density functions (pdfs) are well sampled. Just as the inputs are uncertain so are the outputs of I_s_, Ia and Ic. Thus, stochastic uncertainty modelling of this kind is ubiquitous in almost all risk-related areas of environmental sciences and in many engineering applications, and this has been the case for several decades.

Complexity can be readily added to a BBN network. For example, school size distributions and spatial variation in prevalence can be added to understand how infectious schools are distributed geographically. A transmission model or a model of the presence of siblings in schools can be added. Below we give a single example of calculations for one particular scenario with constant inventory of children and adults in the schools.

Here we reproduce a comprehensive report, generated by UNINET, that records details of the nodes that make up the BBN model underpinning this study, together with one set of mean and quantile results, calculated by stochastic simulation within the program.

### I. General information

1. Bbn file name: E:\Covid-19\Covid BBN\BBN schools Scenario 1 model 260620.uninet.
2. Number of samples per variable: 2000000.

### II. Nodes information

1. Children returning count
  a. Description:
  b. Type: Functional: input data (see Table III).
  c. Dependence info: Child nodes
    - “Infected_Children_count”, functional.
    - “Persons_in_Schools_total”, functional.
2. Infected Children count
  a. Description:
  b. Type: Functional: Children_returning_count*Children_prevalence (see Table III).
  c. Dependence info: Parent nodes
    - “Children_returning_count”, functional.
    - “Children_prevalence”, functional.
  d. Dependence info: Child nodes
    - “Infected_Persons_in_Schools_total”, functional.
3. Teachers total
  a. Description:
  b. Type: Functional: input data (see Table III).
  c. Dependence info: Child nodes
    - “Infected_Teachers_count”, functional.
    - “Persons_in_Schools_total”, functional.
4. Adult prevalence
  a. Description:
  b. Type: Probabilistic: lognormal input data (see Table III).
  c. Dependence info: Child nodes
    - “Infected_Teachers_count”, functional.
    - “Infected_TA_count”, functional.
    - “Infected_Ancillaries_count”, functional.
    - “Children_prevalence”, functional.
5. Infected Teachers count
  a. Description:
  b. Type: Functional: Teachers_return * Adult_prevalence (see Table III).
  c. Dependence info: Parent nodes
    - “Adult_prevalence”, probabilistic.
    - “Teachers_total”, functional.
    - “Teachers_return”, probabilistic.
  d. Dependence info: Child nodes
    - “Infected_Persons_in_Schools_total”, functional.
6. Infected Persons in Schools total
  a. Description:
  b. Type: Functional: Infected_Teachers_count + Infected_Children_count + Infected_TA_count + Infected_Ancillaries_count (see Table III).
  c. Dependence info: Parent nodes
    - “Infected_Children_count”, functional.
    - “Infected_Teachers_count”, functional.
    - “Infected_TA_count”, functional.
    - “Infected_Ancillaries_count”, functional.
  d. Dependence info: Child nodes
    - “Infection_prevalence_in_Schools”, functional.
7. Schools total
  a. Description:
  b. Type: Functional: input data (see Table III).
  c. Dependence info: Child nodes
    - “Persons_per_School_average”, functional.
    - “number_of_Schools_with_infection”, functional.
    - ‘‘percent_Schools_with_infection’’, functional.
8. any School probability zero infections
  a. Description:
  b. Type: Functional: (1- Infection_prevalence_in_Schools)^A^Persons_per_School_average (see Table III).
  c. Dependence info: Parent nodes
    - “Infection_prevalence_in_Schools”, functional.
    - “Persons_per_School_average”, functional.
  d. Dependence info: Child nodes
    - “number_of_Schools_with_infection”, functional.
9. Persons in Schools total
  a. Description:
  b. Type: Functional: Teachers_return + Children_returning_count + TAs_total + Ancillaries_return (see Table III)
  c. Dependence info: Parent nodes
    - “Teachers_total”, functional.
    - “Ancillaries_total”, functional.
    - “Children_returning_count”, functional.
    - “TAs_total”, functional.
    - “Ancillaries_return”, probabilistic.
    - “Teachers_return”, probabilistic.
  d. Dependence info: Child nodes
    - “Infection_prevalence_in_Schools”, functional.
    - “Persons_per_School_average”, functional.
10. Infection prevalence in Schools
  a. Description:
  b. Type: Functional: Infected_Persons_in_Schools_total/Persons_in_Schools_total (see Table III).
  c. Dependence info: Parent nodes
    - “Persons_in_Schools_total”, functional.
    - “Infected_Persons_in_Schools_total”, functional.
  d. Dependence info: Child nodes
    - “any_School_probability_zero_infections”, functional.
11. Persons per School average
  a. Description:
  b. Type: Functional: Persons_in_Schools_total / Schools_total (see Table III).
  c. Dependence info: Parent nodes
    - “Persons_in_Schools_total”, functional.
    - “Schools_total”, functional.
  d. Dependence info: Child nodes
    - ‘‘any_School_probability_zero_infections’’, functional.
12. number of Schools with infection
  a. Description: Binomial(#schools,(1 - any_School_probability_zero_infections’)) is approx Normal with mean = Schools_total*(1- any_School_probability_zero_infections’), variance = any_School_probability_zero_infections*(1- any_School_probability_zero_infections)
  b. Type: Functional: StdNorm_distribution *((1- any_School_probability_zero_infections)* any_School_probability_zero_infections)A0.5 +Schools_total * (1- any_School_probability_zero_infections) (see Table III).
  c. Dependence info: Parent nodes
    - ‘‘any_School_probability_zero_infections’’, functional.
    - “Schools_total”, functional.
    - ‘‘StdNorm_distribution’’, probabilistic.
  d. Dependence info: Child nodes
    - “percent_Shools_with_infection”, functional.
13. percent Schools with infection
  a. Description:
  b. Type: Functional: 100 * number_of_Schools_with_infection/Schools_total (see Table III).
  c. Dependence info: Parent nodes
    - “Schools_total”, functional.
    - “number_of_Schools_with_infection”, functional.
14. Ancillaries total
  a. Description:
  b. Type: Functional: input data (see Table III).
  c. Dependence info: Child nodes
    - “Infected_Ancillaries_count”, functional.
    - “Persons_in_Schools_total”, functional.
15. TAs total
  a. Description: not used
  b. Type: Functional: not used
  c. Dependence info: Child nodes
    - “Infected_TA_count”, functional.
    - “Persons_in_Schools_total”, functional.
16. Infected TA count
  a. Description: not used
  b. Type: Functional: not used
  c. Dependence info: Parent nodes
    - “Adult_prevalence”, probabilistic.
    - “TAs_total”, functional.
  d. Dependence info: Child nodes
    - “Infected_Persons_in_Schools_total”, functional.
17. Infected Ancillaries count
  a. Description:
  b. Type: Functional: Ancillaries_return * Adult_prevalence (see Table III).
  c. Dependence info: Parent nodes
    - “Ancillaries_total”, functional.
    - “Adult_prevalence”, probabilistic.
    - “Ancillaries_return”, probabilistic.
  d. Dependence info: Child nodes
    - “Infected_Persons_in_Schools_total”, functional.
18. Adult child prevalence ratio
  a. Description:
  b. Type: Probabilistic: lognormal (see Table III).
  c. Dependence info: Child nodes
    - ‘‘Children_prevalence’’, functional.
19. Children prevalence
  a. Description:
  b. Type: Functional: Adult_prevalence / Adult_child_prevalence_ratio (see Table III).
  c. Dependence info: Parent nodes
    - ‘‘Adult_child_prevalence_ratio’’, probabilistic.
    - “Adult_prevalence”, probabilistic.
    Dependence info: Child nodes
    - ‘‘Infected_Children_count’’, functional.
20. Ancillaries return
  a. Description:
  b. Type: Probabilistic: Normal distribution (see Table III).
  c. Dependence info: Child nodes
    - “Infected_Ancillaries_count”, functional.
    - “Persons_in_Schools_total”, functional.
21. Teachers return
  a. Description:
  b. Type: Probabilistic: Normal distribution (see Table III).
  c. Dependence info: Child nodes
    - “Persons_in_Schools_total”, functional.
    - “Infected_Teachers_count”, functional.

### III. Simulation values

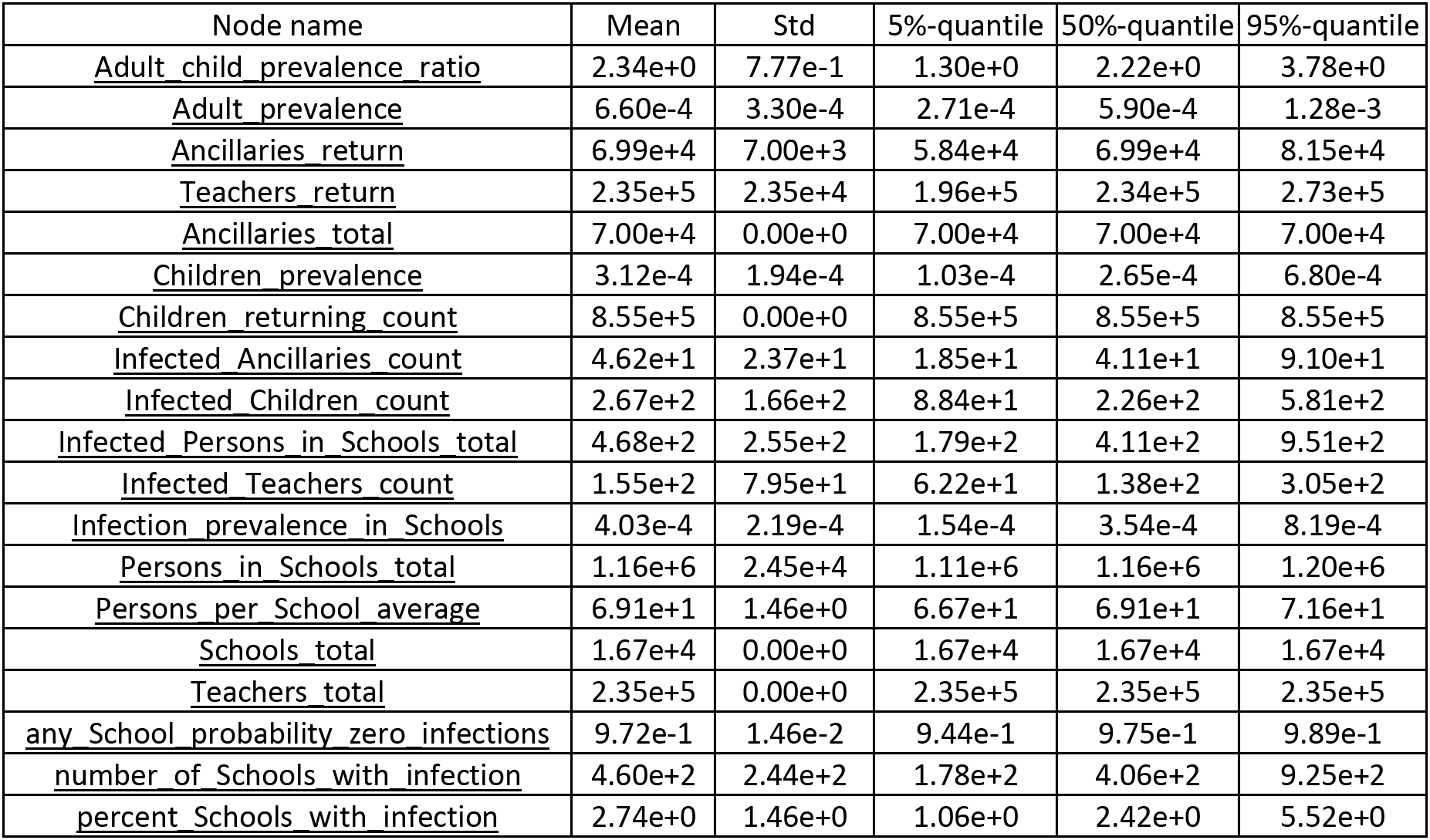

| Node name | Mean | Std | 5%-quantile | 50%-quantile | 95%-quantile |
| --- | --- | --- | --- | --- | --- |
| <u>Adult child prevalence ratio</u> | 2.34e+0 | 7.77e-1 | 1.30e+0 | 2.22e+0 | 3.78e+0 |
| <u>Adult prevalence</u> | 6.60e-4 | 3.30e-4 | 2.71e-4 | 5.90e-4 | 1.28e-3 |
| <u>Ancillaries return</u> | 6.99e+4 | 7.00e+3 | 5.84e+4 | 6.99e+4 | 8.15e+4 |
| <u>Teachers return</u> | 2.35e+5 | 2.35e+4 | 1.96e+5 | 2.34e+5 | 2.73e+5 |
| <u>Ancillaries total</u> | 7.00e+4 | 0.00e+0 | 7.00e+4 | 7.00e+4 | 7.00e+4 |
| <u>Children prevalence</u> | 3.12e-4 | 1.94e-4 | 1.03e-4 | 2.65e-4 | 6.80e-4 |
| <u>Children returning count</u> | 8.55e+5 | 0.00e+0 | 8.55e+5 | 8.55e+5 | 8.55e+5 |
| <u>Infected Ancillaries count</u> | 4.62e+1 | 2.37e+1 | 1.85e+1 | 4.11e+1 | 9.10e+1 |
| <u>Infected Children count</u> | 2.67e+2 | 1.66e+2 | 8.84e+1 | 2.26e+2 | 5.81e+2 |
| <u>Infected Persons in Schools total</u> | 4.68e+2 | 2.55e+2 | 1.79e+2 | 4.11e+2 | 9.51e+2 |
| <u>Infected Teachers count</u> | 1.55e+2 | 7.95e+1 | 6.22e+1 | 1.38e+2 | 3.05e+2 |
| <u>Infection prevalence in Schools</u> | 4.03e-4 | 2.19e-4 | 1.54e-4 | 3.54e-4 | 8.19e-4 |
| <u>Persons in Schools total</u> | 1.16e+6 | 2.45e+4 | 1.11e+6 | 1.16e+6 | 1.20e+6 |
| <u>Persons per School average</u> | 6.91e+1 | 1.46e+0 | 6.67e+1 | 6.91e+1 | 7.16e+1 |
| <u>Schools total</u> | 1.67e+4 | 0.00e+0 | 1.67e+4 | 1.67e+4 | 1.67e+4 |
| <u>Teachers total</u> | 2.35e+5 | 0.00e+0 | 2.35e+5 | 2.35e+5 | 2.35e+5 |
| <u>any School probability zero infections</u> | 9.72e-1 | 1.46e-2 | 9.44e-1 | 9.75e-1 | 9.89e-1 |
| <u>number of Schools with infection</u> | 4.60e+2 | 2.44e+2 | 1.78e+2 | 4.06e+2 | 9.25e+2 |
| <u>percent Schools with infection</u> | 2.74e+0 | 1.46e+0 | 1.06e+0 | 2.42e+0 | 5.52e+0 |

## Appendix 2. Effect of siblings on school infection hazard level estimates

One possible concern in relation to the occurrence of infections in schools is the role of siblings. If a child becomes infectious and has a sibling attending the same school, there is a significantly increased chance that the second sibling would also be infected. We perform a simple scoping calculation to understand the potential impact of co-infected siblings on school infection levels.

Numbers of siblings attending the same state primary school were determined by Nicole and Rabe (2014): they reported that 435,890 siblings in the age group 4 - 16 years lived at same address (including step- and half siblings). To put this in context, according to 2017 data in the ONS National Pupil Database, the total number of pupils aged 5 - 15 years was 7,100,054 pupils; the School Census reported 6,612,292 pupils for the same 2017 cohort. Thus, we adjust Nicole and Rabe’s number by a factor of 10/12 to align it with the ONS age cohort. This leads to an estimated 363,242 sibling pupils in primary schools, or 5.5% of the total number of all pupils.

If we assume an average age difference of 3 years between siblings, then approximately half of all siblings will overlap with their other sibling in a primary school; we conclude that about 2.8% of primary school pupils are siblings attending the same school as their brother or sister. This means 1.4% of all pupils are one-half of a sibling pair. The question then arises, if the probability Sibling 1 is infected is identical with community prevalence, what is the probability Sibling 2 gets infected given their exposure is not an independent sample from a random community process. This probability is needed to compute the effect on infection hazard level due to having two (or more) overlapping, correlated siblings in the same school. Thus, we need to know the likelihood that a second sibling is infected GIVEN his/her sibling is infected.

If cross-sibling infection correlation were to approach 1, the effective prevalence rate for second siblings will be doubled, conditional on the first infection being a random community case. This said, if, as indicated above, only about 1.4% of all pupils are second siblings, with elevated effective prevalence rate, a small marginal increase in mean school infection hazard level is anticipated. Jing et al (2020) suggest sibling infection correlation rate is only about 0.15. Even if this value understates correlated sibling infections in British families, the “sibling effect” only impacts 95th percentile school infection rate estimates in our study.

